# Clinical validation of RCSMS: a rapid and sensitive CRISPR-Cas12a test for the molecular detection of SARS-CoV-2 from saliva

**DOI:** 10.1101/2021.04.26.21256081

**Authors:** Joaquín Abugattás Núñez del Prado, Angélica Quintana Reyes, Juan Blume La Torre, Renzo Gutiérrez Loli, Alejandro Pinzón Olejua, Elena Rocío Chamorro Chirinos, Félix Antonio Loza Mauricio, Jorge L. Maguiña, Julio Leon, Piere Rodríguez Aliaga, Edward Málaga Trillo

**Affiliations:** Facultad de Ciencias y Filosofía, Universidad Peruana Cayetano Heredia, Lima, Perú; Department of Computer Science, Christian-Albrecht University of Kiel, Germany; Hospital Nacional Guillermo Almenara Yrigoyen, EsSalud, Lima, Perú; Hospital Nacional Edgardo Rebagliati Martins, EsSalud, Lima, Perú; Instituto de Evaluación de Tecnologías en Salud e Investigación (IETSI), EsSalud, Lima, Perú; IMS RIKEN Center for Integrative Medical Sciences, Japan; Department of Biology, Stanford University, California, USA

**Keywords:** Perú, saliva test, coronavirus, COVID-19, RCSMS, DETECTR, RT-PCR, SARS-CoV-2

## Abstract

Early detection of SARS-CoV-2 using molecular techniques is paramount to the fight against COVID-19. Due to its high sensitivity and specificity, RT-qPCR is the “gold standard” method for this purpose. However, its technical requirements, processing time and elevated costs hamper its use towards massive and timely molecular testing for COVID-19 in rural and socioeconomically deprived areas of Latin America. The advent and rapid evolution of CRISPR-Cas technology has boosted the development of new pathogen detection methodologies. Recently, DETECTR -a combination of isothermal RT-LAMP amplification and Cas12a-mediated enzymatic detection-has been successfully validated in the Netherlands and the USA as a rapid and low-cost alternative to RT-qPCR for the detection of SARS-CoV-2 from nasopharyngeal swabs. Here, we evaluated the performance of RCSMS, a locally adapted variant of DETECTR, to ascertain the presence of SARS-CoV-2 in saliva samples from 276 patients in two hospitals in Lima, Perú *(current status over a total of 350 samples*). We show that a low-cost thermochemical treatment with TCEP/EDTA is sufficient to inactivate viral particles and cellular nucleases in saliva, eliminating the need to extract viral RNA with commercial kits, as well as the cumbersome nasopharyngeal swab procedure and the requirement of biosafety level 2 laboratories for molecular analyses. Our clinical validation shows that RCSMS detects up to 5 viral copies per reaction in 40 min, with sensitivity and specificity of 93.8% and 99.0% in the field, respectively, relative to RT-qPCR. Since CRISPR-Cas biosensors can be easily reprogrammed by using different guide RNA molecules, RCSMS has the potential to be quickly adapted for the detection of new SARS-CoV-2 variants. Notably, estimation of its negative and positive predictive values suggests that RCSMS can be confidently deployed in both high and low prevalence settings. Furthermore, our field study validates the use of lateral flow strips to easily visualize the presence of SARS-CoV-2, which paves the way to deploy RCSMS as a “point of care” test in environments with limited access to state-of-the-art diagnostic laboratories. In sum, RCSMS is a fast, efficient and inexpensive alternative to RT-qPCR for expanding COVID-19 testing capacity in low- and middle-income countries.

## INTRODUCTION

After sweeping across Asia and Europe, COVID-19 took main stage in Latin America, where socioeconomic disparity and the saturation of public health systems have contributed to alarming infection and mortality rates, particularly in Perú and Brazil. Very conservative estimates from Perú’s National Death Information System (SINADEF) and Ministry of Health (MINSA) show that the impact of COVID-19 on the Peruvian population (∼30 MM) is dramatic, totaling at least 1,730,000 cases, 58,604 deaths and 161,200 excess deaths associated with COVID-19 as of March 30, 2021 (1, 2). The prolonged health emergency has particularly affected vulnerable populations such as indigenous communities, urban and rural areas with low-income, refugees and migrants.

Given the rapid spread of the disease and the arrival of new pandemic waves, it remains urgent to implement early and massive SARS-CoV-2 testing strategies alongside with contact tracing and isolation (3). Molecular tests must be sensitive, rapid and easily accessible to timely identify and treat individuals at high risk of transmitting the infection and efficiently apply partial lockdowns, border closures, and travel restrictions. In addition, molecular testing must be integrated into a genomic surveillance system that tracks the appearance and distribution of viral mutations. In sum, obtaining high-quality diagnostic data is essential to correctly monitor the evolution of the epidemic, ensure the success of public health strategies and the transition to a new normality.

Currently, ∼600 COVID-19 diagnostic kits are commercially offered worldwide (4). These tests are based on the detection of viral genes, viral proteins, or antibodies generated by the human immune system against the virus. While antibody tests are recommended for seroprevalence and epidemiological studies, the “gold standard” among COVID-19 molecular tests is the quantitative reverse transcription-polymerase chain reaction (RT-qPCR) (5). Recently, viral antigen detection tests have added speed and accessibility to COVID-19 molecular testing, although their lower sensitivity relative to RT-qPCR limits their utility to individuals with high viral loads and risk of transmission (6). In developing countries, RT-qPCR testing at the population level is restricted by poor access to adequate equipment, supplies and infrastructure, as well as by the need for trained personnel and high biosafety standards; in fact, most diagnostic kits in Latin America are produced in the developed world (7). Hence, the local development and distribution of molecular detection methods are key to the fight against COVID-19 in these countries.

DETECTR is a recently developed molecular test that uses the Nobel-prize winning CRISPR-Cas technology to detect SARS-CoV-2 RNA extracted from nasopharyngeal swabs, with high efficiency (8). This method couples two reactions: i) the first reaction performs simultaneous reverse transcription and loop-mediated isothermal amplification (RT–LAMP) to generate DNA copies from viral RNA and amplify them exponentially at a single constant temperature (between 60 and 65°C), using three pairs of nested primers and a DNA polymerase with displacement and replication activities (9), and ii) the second reaction exploits the CRISPR-Cas12a recognition system to target and detect the RT-LAMP-amplified viral gene sequence, unleashing the collateral DNAse activity of Cas12a and leading to the indiscriminate cut of single-stranded DNA reporter molecules that produce either fluorescent or immunochromatographic readouts upon cleavage (8). A significant advantage of DETECTR over other methods is that it requires neither sophisticated equipment nor specialized personnel, and it is a low-cost alternative with sensitivity and specificity comparable to RT-qPCR (6). Altogether, RT-LAMP and CRISPR-Cas12a reactions have a processing time of ∼40 minutes, very convenient compared to the 2-4 hours required for the RT-qPCR reaction and its even longer data processing time (6). Another important advantage of DETECTR over RT-qPCR is its ability to confirm the presence of SARS-CoV-2 RNA not only via standard fluorescence but also by using lateral flow strips, which simply requires replacing the fluorophore quencher with biotin in the reporter molecules. This results in an inexpensive visualization format, similar to a pregnancy test, easy to apply and interpret (8). Importantly, various molecular tests that similarly combine RT-LAMP and CRISPR-Cas have successfully been used to detect Zika, Dengue and HIV viruses in humans, and some coronaviruses in animals (10).

COVID-19 Nucleic Acid Tests *(NATs*) -such as DETECTR or RT-qPCR-have two major limitations related to sample collection. On one hand, the of nasopharyngeal swab procedure is uncomfortable for patients, risky for sample collection personnel, and generates the need for swabs and costly viral transport media. On the other hand, the extraction of high-quality viral RNA in standard laboratories requires commercial kits, expensive reagents and long processing times. Interestingly, recent studies have shown that respiratory samples can be directly lysed and used efficiently for molecular diagnostics without commercial RNA extraction kits (11-14). This simple incubation method uses standard low-cost laboratory equipment and is based on a simple thermochemical reaction that guarantees 1) inactivation of RNases in the sample and 2) lysis of viral particles, thus ensuring the release of viral genetic material and rendering the sample non-infectious (11-14). This is of great relevance for COVID-19 molecular testing in low and middle-income countries because it eliminates the need for highly trained personnel and biosafety level 2 facilities.

In the present study, we report the development, standardization and clinical validation of a new procedure that combines the efficiency and robustness of DETECTR (i.e. RT-LAMP and CRISPR-Cas coupled reactions) with the rapid and low-cost TCEP/EDTA thermochemical method for viral RNA preparation from saliva samples. We show that our integrated system -called *Rapid Coronavirus-Sensitive Monitoring from Saliva, RCSMS*-successfully detected SARS-CoV-2 RNA in a set of 276 saliva samples (*current status from a total of 350 samples*) from patients from two hospitals in Lima. The clinical diagnostic performance of RCSMS was validated by comparison with routine RT-qPCR from nasopharyngeal swabs under field conditions. Together, our results show that RCSMS is a simple and easy-to-implement molecular test at the primary care level, outside the laboratory, with a great potential to contribute to the massification of COVID-19 molecular testing in Peru and neighboring countries.

## MATERIALS AND METHODS

### Study design

The present study, aimed at validating the use of RCMS to test for the presence of SARS-CoV-2 in saliva from Peruvian patients, was developed in three stages:

#### 1) Method optimization

To establish the appropriate experimental conditions for the molecular detection of SARS-CoV-2 via DETECTR, synthetic viral RNA was generated from DNA templates and taken as input material (see below), using PCR products as amplification control for DETECTR. This stage evaluated the performance of DETECTR under ideal conditions, in particular: a) the robustness of the RT-LAMP reaction upon changes in reaction time (between 20 and 30 min) and temperature (gradient between 62 and 68°C), b) the limit of detection (LOD) of the CRISPR-Cas reaction (serial dilutions of the RNA substrate for RT-LAMP), c) the robustness of the CRISPR-Cas reaction to changes in the final concentration of Cas12a enzyme and guide RNAs (0.5, 1, 2 and 3X) in the ribonucleoprotein complex (RNP), length of guide RNAs (41 and 44 bp), preincubation time for the formation of the RNP complex (10, 20 and 30 min), length of fluorescent and biotinylated ssDNA probes (5 and 8 bp), final concentration of fluorescent (20, 50 and 100 nM) and biotinylated probes (20, 50, 100, 200, 400, 500 and 600 nM), CRISPR detection time by fluorescence (5-30 min) and immunochromatography (10, 20 and 30 min), amount of RT-LAMP product (1, 2, 3 and 5 µl) and d) the repeatability of measurements (three replicas). These evaluations also allowed us to optimize the efficiency, cost and readout levels of the test, as well as to verify its reproducibility upon variations in instrumentation and operators.

#### 2) Analytical validation

To assess the analytical specificity of DETECTR, we relied on *in vitro* data from a similar study (8) and examined the possibility of cross-reactivity in the primers and guide RNAs used here. For this, we ran a comparative *in silico* analysis of the corresponding regions in common human coronavirus sequences (HCoV-HKU1 (NC_006577.2), HCoV-NL63 (NC_005831.2), HCoV-OC43 strain ATCC VR-759 (NC_006213.1), MERS-CoV (NC_019843.3), SARS-CoV (NC_004718) and SARS-CoV-2; NC_045512). The potential ability of the test to detect viral variants currently circulating in Perú was inferred by aligning the sequences of our DNA primers and guide RNAs with those of 944 viral genomes from Peruvian patients (https://nextstrain.org/community/quipupe/Nextstrain_Peru and https://www.gisaid.org/, accessed March 31, 2021). To evaluate the performance of DETECTR on Peruvian samples under optimal laboratory conditions, we followed the criteria established by the Foundation for Innovative New Diagnostics (FIND, https://www.finddx.org/covid-19/sarscov2-eval/). The retrospective analysis was carried out on 100 anonymized nasopharyngeal swabs with recorded negative (n=50) and positive (n=50) results by RT-qPCR. The samples, kindly provided by the Peruvian National Institute of Health (INS), which lacked medical records, were obtained by the INS during epidemiological evaluations between March and June 2020, and remained stored in their laboratories until they were delivered to our team on June 30, 2020. Once received at 4°C, the 100 samples were analyzed via DETECTR for E and N viral genes, with two additional (blinded and randomized) repetitions, to determine sensitivity and specificity values, as well as repeatability and limit of detection (analytical sensitivity).

#### 3) Clinical Validation

The performance of RCSMS under real field conditions was evaluated by means of a prospective cross-sectional, observational study of diagnostic test precision. To date, samples from 276 study subjects (out of a total of 350) from Guillermo Almenara Irigoyen and Edgardo Rebagliati Martins National Hospitals have been tested following a consecutive non-probabilistic sampling scheme. The sample size was determined using the sensitivity and specificity estimation formula in the PASS 11.0 program (NCSS, LLC. Kaysville, Utah, USA), considering an expected sensitivity and specificity of 96% and 99%, respectively, as reported by Mayuramart et al. (15), and assuming a confidence level of 95% and a precision of 5%. Given the lack of information about the positivity rate in these hospitals, equal proportions (50%/50%) were assumed for the presence or absence of SARS-CoV-2 infection in these patients, as suggested by Macfarlane (16). Regarding eligibility criteria, we selected ambulatory patients over 18 years of age who visited the hospital for SARS-CoV-2 detection having reported clinical symptomatology defined as: a) *acute respiratory infection*: cough and/or sore throat, general malaise, fever, headache, nasal congestion, shortness of breath, loss of smell, loss of taste, or b) *severe acute respiratory infection*: temperature ≥ 38°C and cough, with onset within the last 10 days. Subjects who had participated in vaccine clinical trials of vaccination programs against SARS-CoV-2 were excluded from the study, as were subjects with xerostomia, users of antiparkinsonian drugs, antipsychotics, and/or neuroleptics, and patients with more than three weeks of illness counted since the onset of symptoms. Likewise, saliva samples of poor quality (containing blood, food residues or phlegm) and volumes outside the optimal range (1-2 ml) were excluded. Upon being instructed about the study, the patients read and signed the informed consent. Nasopharyngeal swabs and saliva samples were collected following WHO recommendations, and molecularly analyzed in parallel (see below). For each participant, an absolute code was assigned to the data collection form and the samples, to protect their personal data and maintain the confidentiality of the results. Saliva samples were transferred the same day of the collection at 4°C to UPCH for RCSMS analysis of the E gene; nasopharyngeal swab samples remained at Guillermo Almenara Irigoyen National Hospital to be analyzed by RT-qPCR. Handling of the nasopharyngeal swabs and saliva samples was conducted under recommended biosafety standards for respiratory viruses. The analyses were run blind, so that the personnel in each laboratory ignored of test results of the other laboratory. Finally, we also evaluated the LOD of RCSMS by testing serial dilutions of synthetic RNA in a range of 5,000 to 1 viral copy per 10 µl RT-LAMP reaction, which included 2 µl of saliva inactivated with TCEP / EDTA and negative for SARS-CoV-2.

### Generation of synthetic viral RNA templates by PCR and in vitro transcription

To select the ideal detection targets for this study as well as to establish their optimal amplification and detection parameters, we generated a panel of synthetic RNA templates encoding the viral genes N (934 bp), E (532 bp), S (540 bp), from Nsp6 to Nsp8 (734 bp), from Nsp10 to Nsp12 (798 bp) and the human POP7 gene (406 bp, sample quality control). These RNA templates were generated via *in vitro* transcription from PCR products modified to contain the sequence of the T7 transcriptional promoter at their 5 ’end to allow their use *in vitro* transcription templates. The PCR reactions were performed using GoTaq G2 DNA polymerase (Promega) according to the conditions and cycling profile shown in Table 2. After resolving the amplification products on 1% agarose gels, the corresponding bands were cut and purified with help of the GeneJet Gel DNA Extraction Kit (ThermoScientific). The DNA concentration in the eluates was quantified using a Nanodrop 2000 spectrophotometer (ThermoScientific) and 100 ng DNA were used for the in vitro transcription reaction using the AmpliScribe T7-Flash Transcription Kit (Lucigen), following the manufacturer’s recommendations. The resulting RNA templates were purified with the GenElute kit (Sigma) and quantified with the Qubit 4.0 fluorometer using the RNA BR assay kit (Invitrogen). In addition, the integrity of the RNA was verified by denaturing agarose electrophoresis.

### Inactivation of saliva samples and RNA extraction

Saliva samples containing 0.01 volumes of the 100X inactivation solution (0.25 M TCEP-HCl, 0.1 M EDTA and 1.15 N NaOH) were mixed and homogenized with a vortex for 10-15 sec, then heated for 10 min at 95°C and cooled on ice. For extraction controls from saliva and nasopharyngeal swab samples, RNA was extracted using the ReliaPrep Viral TNA Miniprep Kit, Custom (Promega) and the QIAamp Viral RNA Mini Kit (Qiagen).

### RT-LAMP amplification

The sets of six primers for the LAMP amplification of each viral gene (Table 1) were previously conjugated in a 10X master mix (Table 3). For the RT-LAMP reaction, a mixture of 0.2 µl Warmstart RTx Reverse Transcriptase and 0.5 µl Bst 3.0 DNA polymerase (NEB) were used, taking as input material 5 µl of purified RNA or 2 µl of inactivated saliva in a total volume of 10 µl (8 mM final concentration of MgSO4) and running the reactions for 30 min at 62°C.

**Table 1:**
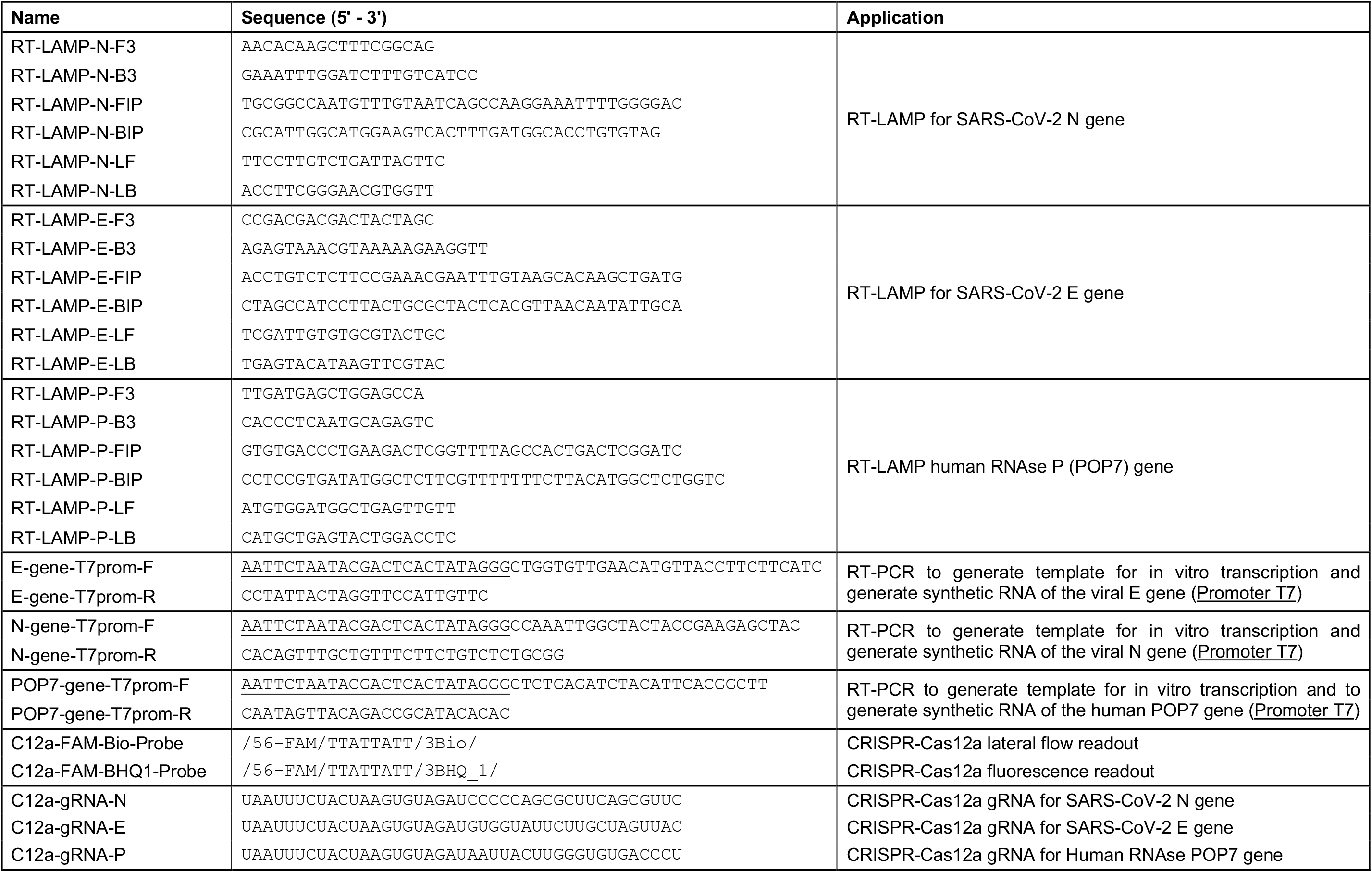
Sequences of primers, guide RNAs and probes used in the study:

**Table 2:** Recipe and program of the thermal cycler for the generation of viral DNA templates by RT-PCR, modified with the T7 promoter for in vitro transcription of positive control RNAs:

| Component | Initial concentration | Final concentration | Volume (µl) |
| --- | --- | --- | --- |
| H<sub>2</sub>O | -- | -- | 5.12 µl |
| 5X Buffer | 5X | 1X | 3 µl |
| dNTPs | 10 mM | 0.2 mM | 0.3 µl |
| Primer 1 | 10 mM | 0.33 mM | 0.5 µl |
| Primer 2 | 10 mM | 0.33 mM | 0.5 µl |
| GoTaq G2 DNA Polymerase | 5 U/µl | 0.027 U/µl | 0.08 µl |
| M-MLV Reverse Transcriptase | 200 U/µl | 6.67 U/µl | 0.5 µl |
| Mastermix volume per reaction |  |  | 10 µl |
| pCCI-4K-SARS-CoV-2-Wuhan-Hu-1 |  |  | 5 µl (5 ng) |
| Total volumen per reaction |  |  | 15 µl |

| Step | Temperatura (°C) | Tiempo (s) |
| --- | --- | --- |
| Reverse Transcription | 45 | 1200 |
| Initial Denat. | 94 | 120 |
| 35X Cycles | 94 | 30 |
|  | 60 | 30 |
|  | 72 | 60 |
| Final Ext. | 72 | 300 |

**Table 3:** Proportions for Master Mix (10X) for RT-LAMP primers.

| LAMP Primer (100uM en H2O) | Final concentration | Volume ( µl) |
| --- | --- | --- |
| F3 | 2 µM | 20 µl |
| B3 | 2 µM | 20 µl |
| FIP | 16 µM | 160 µl |
| BIP | 16 µM | 160 µl |
| LF | 8 µM | 80 µl |
| LB | 8 µM | 80 µl |
| Nuclease free water | - | 480 µl |
| Total Volume |  | 1000 µl |

### CRISPR-Cas detection

For the formation of the ribonucleoprotein complex (RNP = Cas12a + guide RNA), the LbCas12a enzyme (NEB, 50 nM final concentration) was incubated with the guide RNA for each viral target (62.5 nM, final concentration) in 1X NEBuffer 2.1 for 30 min at 37°C. ssDNA probes (Table 1) were added to the mix at a concentration of 20 nM for fluorescence measurements (485 nm excitation, 528 nm emission) in a Cytation 7 plate reader (BioTek) 500 nM for lateral flow strips (Milenia GenLine Hybridetect, Milenia Biotech). For each fluorescence reaction, 2 µl of RT-LAMP product, 80 µl of 1X NEBuffer 2.1 and 20 µl of RNP were mixed per well of a 96-well plate (Costar). For lateral flow strips, 2 µl of RT-LAMP product and 20 µl of RNP were mixed and incubated in a 1.5 ml tube for 10 min at 37°C, and then 80 µl of pre-warmed Milenia GenLine Dipstick Assay Buffer buffer were added (for more details: https://www.milenia-biotec.com/en/tips-lateral-flow-readouts-crispr-cas-strategies/). For the sequences of all guide RNAs and CRISPR ssDNA probes, see Table 1. The positivity threshold was calculated as the ratio between signal and minimum fluorescence values (“fold change”) detected after 10 min of measurement. The reading and interpretation of the results in lateral flow strips is based on the appearance of diagnostic bands (Fig. 1 and Table 4), while for fluorescence results, real-time or end-point measurements were made via automated reading on 96-well plates.

**Table 4:** Interpretation of results by immunochromatographic lateral flow strips. For reference on reading the lateral flow strips, see Figure 1.

| Gene N o E | Gene P | Interpretation |
| --- | --- | --- |
| + | +/- | Positive Result for SARS-CoV 2 |
| - | + | Negative Result for SARS-CoV 2 |
| - | - | Invalid Result |

**Figure 1.**
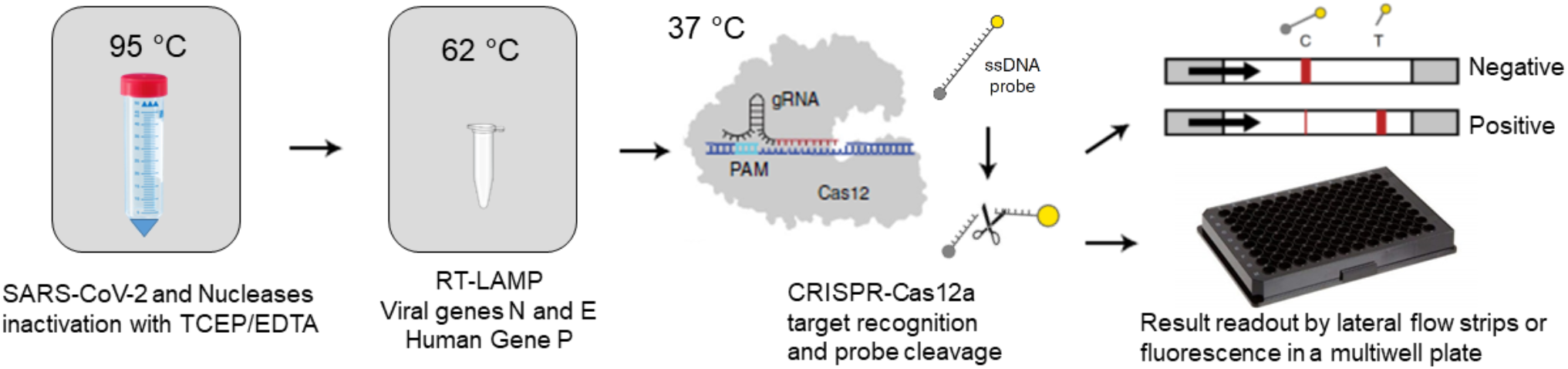
RCSMS detection workflow. Upon saliva treatment with TCEP/EDTA, 2 µl of inactivated sample are added to a 10 µl RT-LAMP reaction. A 2 µl aliquot of the RT-LAMP product is then mixed with the RNP complex consisting of Cas12 and RNA guides. Recognition of viral target sequences by the RNP complex triggers the collateral activity of Cas12a, resulting in the cleavage of ssDNA reporter probes. For immunocromatographyic (qualitative) readout, a lateral flow strip is then inserted into the CRISPR-Cas12a reaction tube or well. Within two minutes, uncleaved reporter molecules flow and accumulate into the control capture line of the strip (C band in the image), whereas cleaved reporter molecules flow towards the target capture line of the strip (T band in the image), (adapted from Broughton et al. (8) and Patchsung et al. (18)). For fluorescence (quantitavite) readout, CRISPR-Cas12a reactions are recorded in real-time over 10 min using an automated plater reader; cleaved reporter molecules yield a bright fluorescent signal.

### Amplification and detection by RT-qPCR

The RT-qPCR of viral genes Orf1ab and N run at UPCH used the *in vitro* diagnostic (IVD) FTD-SARS-CoV-2 kit (Siemens) and QTower 3G real-time thermocyclers (Analytik-Jena). The RT-qPCRs performed at the Guillermo Almenara Irigoyen National Hospital used the IVD Viasure SARS-CoV-2 Real Time PCR Detection Kit (CerTest Biotec), which also amplifies the viral genes Orf1ab and N and the Rotor-Gene real-time PCR cycler (Qiagen). Diagnostic Ct threshold were set as recommended by each kit and according to WHO guidelines.

### Statistical analysis

A descriptive analysis of the sociodemographic and clinical characteristics of the population was carried out. These included measures of central tendency and dispersion that were calculated for the quantitative variables, as well as absolute and relative frequencies for the categorical variables. Likewise, the level of concordance (Cohen’s Kappa coefficient) between RT-qPCR and RCSMS was calculated for both analytical and clinical validations, as well as sensitivity, specificity, positive predictive value (PPV), negative predictive value (NPV), the area under the curve (AUC), accuracy of RCSMS, and PPV/ NPV simulations assuming various prevalence scenarios. A stratified analysis by time was also included (greater than 7 and less than 7 days of illness). The estimates included 95% confidence intervals (95% CI) with an exact binomial method, and a p value <0.05 was considered significant. All analyses were performed with the Stata software version 16.0 (StataCorp, College Station, TX, USA). For data visualization and graphs GraphPad Prism 7.00 was used (GraphPad Software, La Jolla California USA, www.graphpad.com).

### Ethical considerations

Participants in the clinical validation stage were enrolled and sampled according to the ethics protocols approved by the INCOR EsSalud CEI (certificate 02/2021-CEI) and UPCH (SIDISI code 202099). All the participants read and signed informed consents prior to providing samples for this study.

## RESULTS

### Amplification and Detection of SARS-CoV-2 sequences by DETECTR

Synthetic RNA fragments of the viral genes N (934 bp) and E (540 bp), and the human POP7 gene (406 bp), were selected and used as templates to optimize the conditions for RT-LAMP reactions (Fig. 2A and B). The RT and LAMP reactions were run simultaneously in a single tube, taking 10,000 copies of synthetic RNA fragments as starting material, and using reaction time and temperature gradients. Comparative evaluation of the reactions by real-time fluorescence and agarose gel electrophoresis revealed that optimal amplification of the three fragments occurred after 30 minutes at 62°C (Fig. 2C).

**Figure 2.**
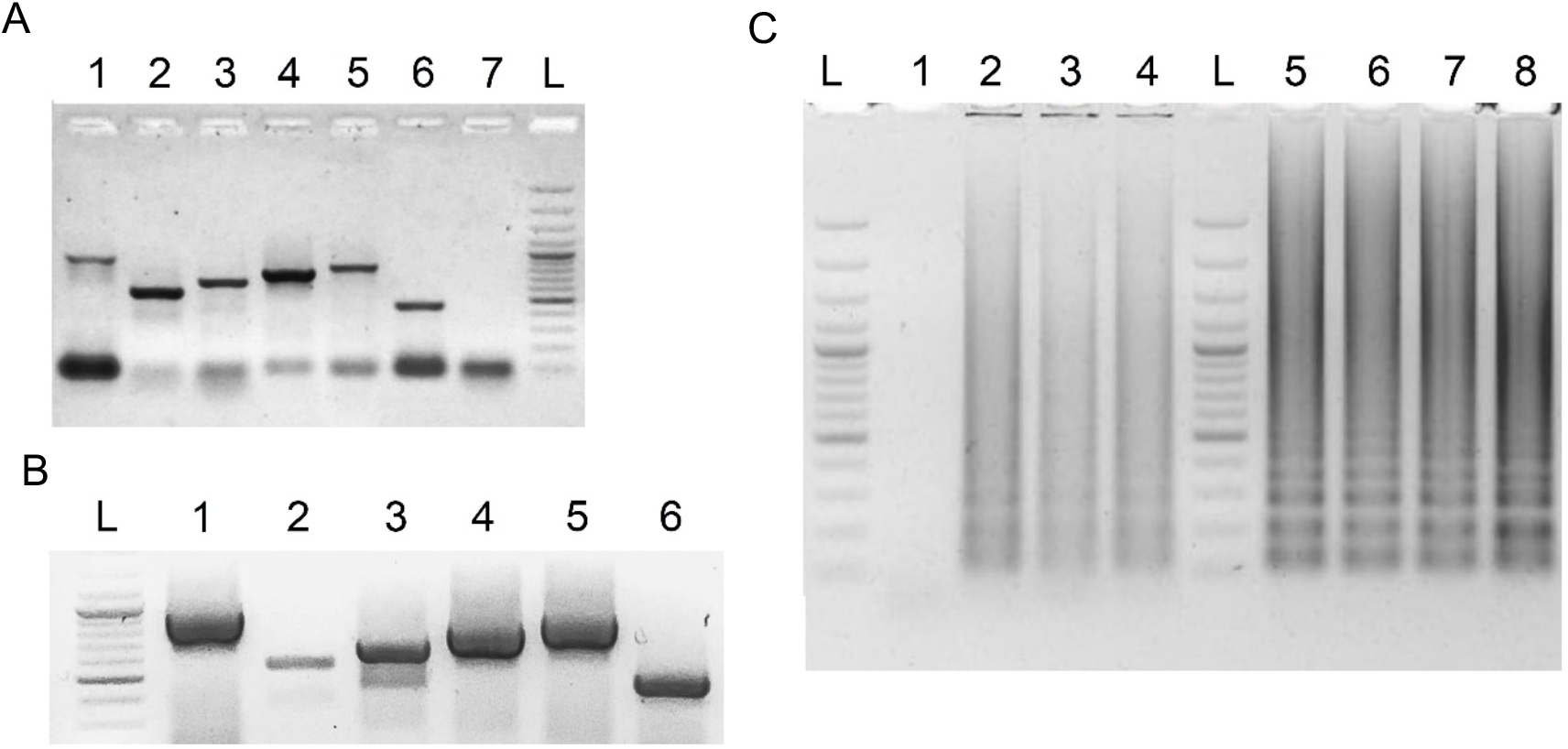
PCR and RT-LAMP templates and products. Visualization of various RT-PCR products, *in vitro* synthetized RNA templates and RT-LAMP products used for RCSMS standardization, on 1% agarose gels stained with ethidium bromide. (A) RT-PCR products used as DNA templates for the generation of *in vitro* transcribed RNAs; Lane 1: 931 bp viral N gene fragment, Lane 2: 557 bp viral E gene fragment, Lane 3: 653 bp viral S gene fragment; Lane 4: 751 bp viral Nsp6 to Nsp8 gene fragment, Lane 5: 815 bp viral Nsp10 to Nsp12 gene fragment, Lane 6: 428 bp human POP7 RNAse gene fragment, Lane 7: RT-PCR Negative control, Lane L: GenRuler 100bp Plus DNA Ladder (Thermo). (B) *in vitro* generated RNA templates; Lane 1: viral N gene; Lane 2: viral E gene; Lane 3: viral S gene; Lane 4: viral Nsp6 to Nsp8 gene; Lane 5: viral Nsp10 to Nsp12 gene; Lane 6: human RNAse POP7 gene. (C) Titration of E gene RNA input in 10 µl RT-LAMP reactions to obtain the characteristic LAMP ladder product; Lane L GenRuler 100bp Plus DNA Ladder (Thermo); Lane 1: reaction with 0 copies; Lane 2: reaction with 1 copy; Lane 3: reaction with 5 copies, Lane 4: reaction with 10 copies; Lane 5: reaction with 20 copies; Lane 6: reaction with 50; Lane 7: reaction with 100 copies; Lane 8: reaction with 250 copies.

The next step involved the detection of the amplified viral fragments using CRISPR-Cas technology. To this end, we used 2 µl of the RT-LAMP amplified products as substrate and 2 µl of RT-PCR products -generated from the same synthetic templates-as positive controls. The minimum and optimal pre-incubation time for the formation of the RNP complex (i.e. Cas12a + guide RNA) was 10 min but it could be extended up to 30 min. Once formed, RNPs could be stored for up to 48 hours at 4°C. The detection reactions for the E, N and POP7 genes worked optimally when run for 10 minutes at 37°C in the presence of 50 nM of RNP complex, formed by equimolar concentrations of Cas12a enzyme and RNA guides, regardless of the length of the latter. 8 bp reporter probes yielded optimal signals at 20 nM (fluorescent readout) and 500 nM (immunochromatography readout) (Fig. 3 and See Supplementary Material) The minimum threshold of positivity of the fluorescence measurements was set at 20,000 Relative Fluorescence Units (UFR) after 10 min of reaction, and with a maximum of 100,000 UFR. On the other hand, the appearance of immunochromatographic signals on the lateral flow strips was consistently achieved after 2 min incubation of the strips in the reaction tubes. The limit of detection (LOD) for both viral genes was determined using serial dilutions of the synthetic RNA templates, covering the range from 250 to 1 viral copy per 10 µl RT-LAMP reaction. Remarkably, our assay was able of detect down to 1 copy of both N and E genes in fluorescence and immunochromatographic readings (Fig. 4A and B and Table 5), showing excellent concordance and reproducibility between both detection formats.

**Table 5:** Limit of Detection (LOD) assay for analytical and clinical validation. RT-LAMP reactions of 10 µl carried out in triplicate with synthetic RNA of the viral genes E and N, were used as substrate for CRISPR-Cas12a detection. Up to 1 viral copy per reaction was detected in both genes during analytical validation and up to 5 viral copies per reaction (including 2 µl of SARS-CoV-2 negative inactivated saliva) during clinical validation.

| Copies / reaction | Analytical Validation |  |  |  |  |  | Copies / reaction | Clinical validation |  |  |
| --- | --- | --- | --- | --- | --- | --- | --- | --- | --- | --- |
|  | Gene E |  |  | Gene N |  |  |  | Gene E |  |  |
| 250 | + | + | + | + | + | + | 5000 | + | + | + |
| 100 | + | + | + | + | + | + | 500 | + | + | + |
| 50 | + | + | + | + | + | + | 50 | + | + | + |
| 20 | + | + | + | + | + | + | 5 | - | + | - |
| 10 | + | + | + | + | + | + | 1 | - | - | - |
| 5 | + | + | + | + | + | + | 0 | - | - | - |
| 1 | + | + | + | + | + | + |  |  |  |  |
| 0 | - | - | - | - | - | - |  |  |  |  |

**Figure 3.**
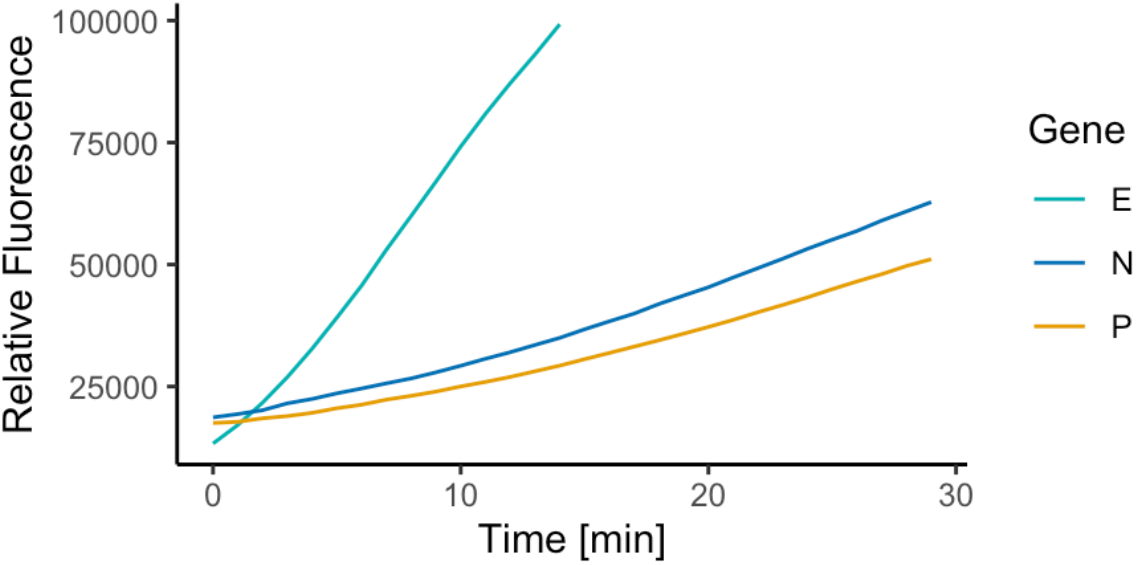
Fluorescent CRISPR-Cas12a-based detection of SARS-CoV-2 genes. Synthetic RNAs corresponding to the viral E, N and human POP7 genes were used as templates for RT-LAMP. The amplification products were then used as substrates for CRISPR-Cas12a reactions in the presence of fluorescent ssDNA reporter probes (485 nm excitation, 528 nm emission). Real-time fluorescence measurements of CRISPR-Cas12a reactions were performed separately for each gene, routinely every minute for 10 min at 37°C under the green channel of an automated plate reader. Signals obtained at every time point were normalized against the background signal detected at t=o; the curves represent the change in normalized fluorescence units (I-I_0_, y axis) over time (min, x axis). In the example shown, the red, green and yellow curves correspond to 30 min data collection for N, E and P genes, respectively. During such extended measurements, the E gene signal reached the instrument’s top detection limit after 15 min, whereas the N and P gene signals continued to grow at a lower rate.

**Figure 4.**
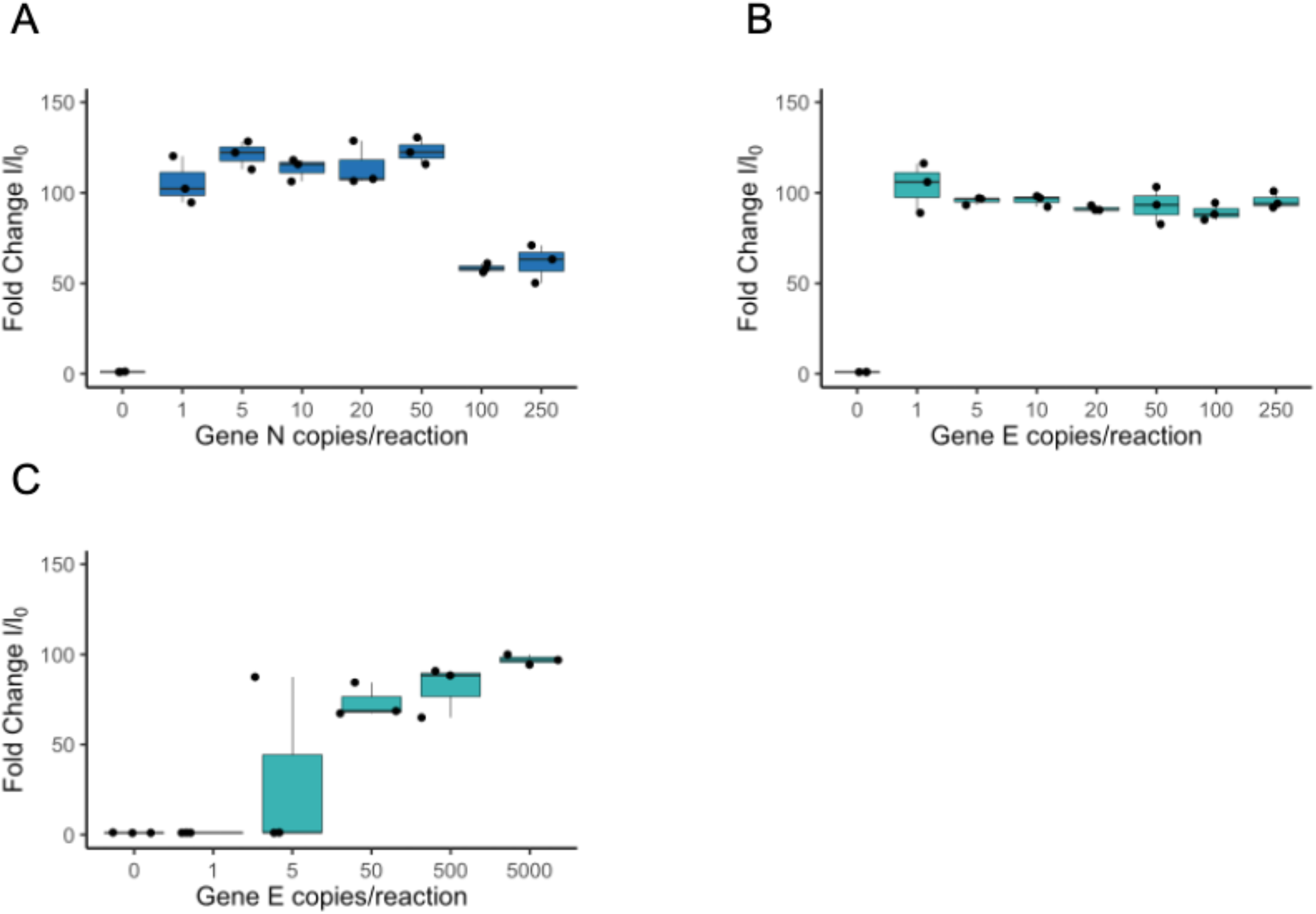
LOD estimations for analytical and clinical validations. Limits of detection were estimated from fluorescent CRISPR-Cas12a assays under laboratory and field conditions (lateral flow assays not shown here). The corresponding RT-LAMP reactions were performed in triplicate using serial dilutions of synthetic RNA templates in water or saliva. The graphs depict the fold change in fluorescence (y axis) vs the number of viral copies per RT-LAMP reaction (x axis). Fold change values (I/I_0_) were obtained by normalizing the fluorescent signal of the sample end-point (t=10 min) against the background signal at t=0. The lines between the dots represent the arithmetic mean with standard deviation for the three samples run for any given value of viral copies/reaction. (A) and (B) In an optimal laboratory setting, down to 1 viral copy per 10 µl RT-LAMP reaction was detected for both N and E genes. (C) Under field conditions down to 5 viral copies per 10 µl RT-LAMP reaction were detected.

### Analytical validation of DETECTR in Peruvian samples

To rule out the possibility of cross-detecting other, related coronaviruses in the sample, we performed an *in silico* analysis of the RT-LAMP primers and guide RNAs used in this study, by aligning their nucleotide sequences with the corresponding regions in the viral E and N genes of different human coronaviruses (see Materials and Methods). The analysis did not show significant similarities, except for the F3 primer of the E gene, which is incapable of producing amplification by itself, thus excluding its cross-reactivity (see Discussion).

The appearance of SARS-CoV-2 variants worldwide makes it necessary to update all diagnostic and therapeutic methods based on the molecular recognition of viral genes and proteins. Therefore, we analyzed the 944 Peruvian genomes available in the Nextstrain and GISAID databases by March 31, 2021, and examined all nucleotide polymorphisms within the E and N genes, which are the targets of this study (see Materials and Methods). The analysis confirmed the high conservation of the viral genomes and the absence of mutational hot spots in the N and E genes corresponding to the binding sites of primers and guide RNA. However, we identified a few minor exceptions (see Table 6). For example, for the E gene, we identified two unique viral sequences containing each a C→T transition (singleton mutations) at different positions of the F3 primer-binding site. For the N gene, a singleton mutation was found at the guide RNA binding site, as well as four mutations at the primer B3 binding site. The first two, singletons, and the third one (found only in two viral sequences) are located at the 5 ’end of the target region. The fourth mutation, close to the 3 ’end of the target region, was found in 22 sequences. None of these point mutations, however, disrupted the amplification efficiency of either E or N genes by RT-LAMP (see Discussion).

**Table 6:**
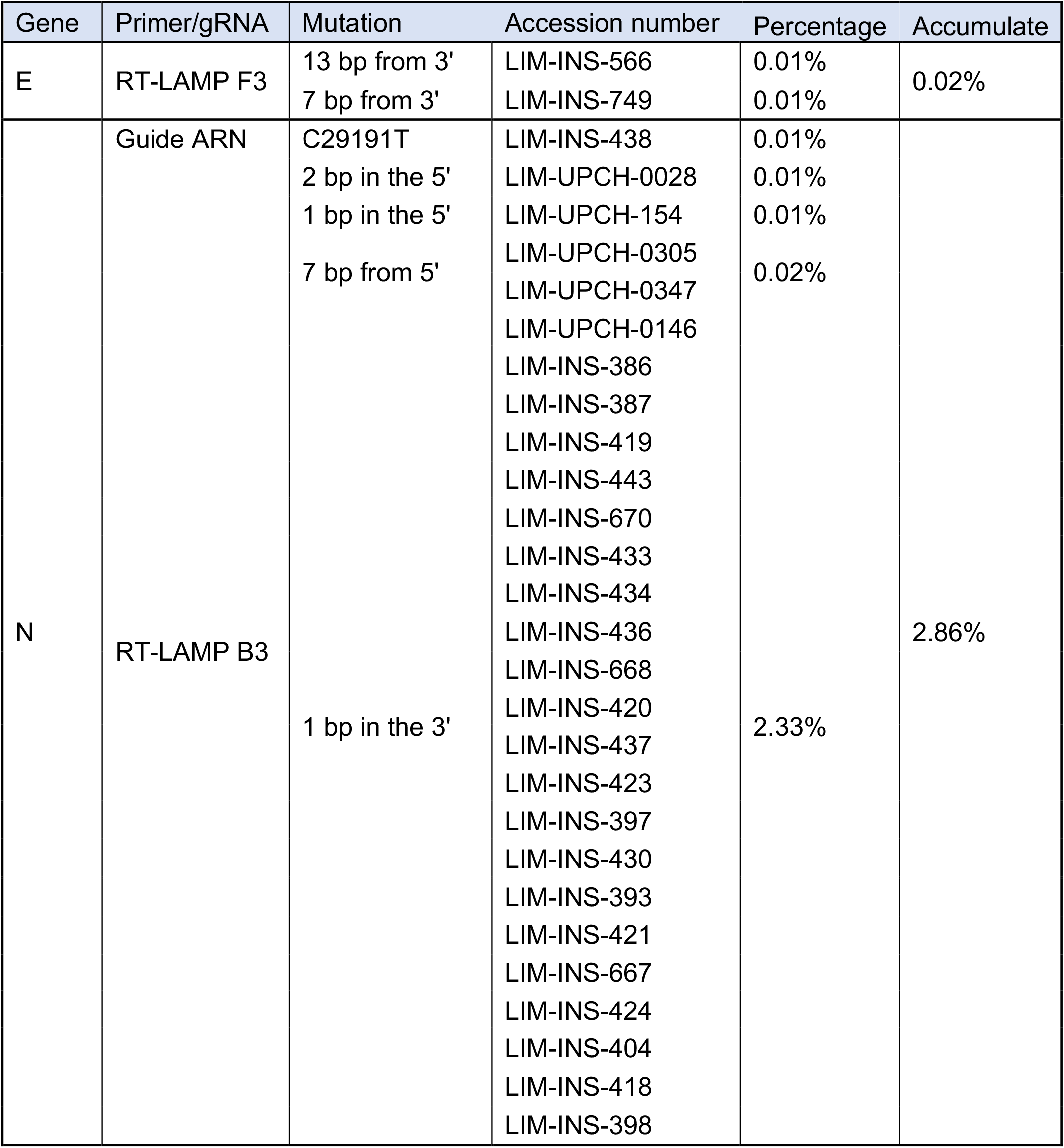
Mutations in Peruvian sequences matching guide RNA binding sites for Cas12a and RT-LAMP primers.

For the retrospective analytical validation, we used nasal swabs because saliva samples were not available at the official repository of SARS-CoV-2 samples in Perú (INS). Viral RNA was extracted from these clinical samples by conventional commercial methods and used as template for the amplification and detection of the viral genes E and N via DETECTR. Owing to the absence of clinical and epidemiological data from the 100 nasal swabs, as well as to the possibility of having undergone RNA degradation during storage/transportation, we introduced an additional verification step by RT-qPCR (Orf1ab and N genes) as an internal control. Notably, both DETECTR assays with fluorescence or immunochromatography readouts and RT-qPCR yielded 39 positive and 59 negative results for SARS-CoV-2, plus two negative samples for the human RNase P gene (Fig. 5), which deviates from the expected number of 50 positive and 50 negative samples indicated at the moment we received the samples. This inconsistency likely stems from the partial degradation of viral RNA in 11 positive samples and the degradation of total RNA in two negative samples, possibly caused by the storage conditions and the transfer of the samples at 4°C. Similar inconsistencies were independently reported by other groups who received similar samples from INS to validate their own diagnostic kits. Moreover, three rounds of analyses reproduced these results, with only two positive samples yielding a negative result in one of their three replicates. Therefore, the corrected values for positivity and negativity actually amount to 39.8% and 60.2%, respectively. The Kappa-Cohen index of 1.0 (95% CI: 1.0-1.0; p <0.00001) reaffirms the total agreement between the two detection methodologies (DETECTR and RT-qPCR), with sensitivity values of 100% (95% CI: 91.0-100.0) and specificity of 100% (95% CI: 94.0-100.0), as well as a positive predictive value (PPV) of 100% (95% CI: 91.0-100.0) and a negative predictive value (NPV) of 100% (95% CI: 94.0-100.0) (Table 7 and Table 9). Finally, as a measure of performance, the Area Under the Curve (AUC) of the Receiver Operating Characteristic (ROC) curve was calculated at 1.0 (95% CI: 1.0-1.0).

**Table 7:**
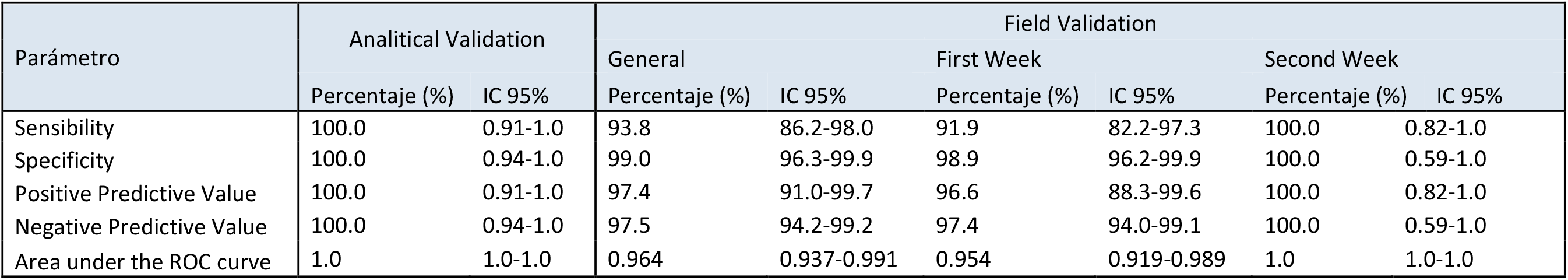
Diagnostic performance measures of RT-LAMP / CRISPR-Cas in analytical and clinical validation. Measures of sensitivity, specificity, positive predictive value, negative predictive value, accuracy and area under the ROC curve of the RT-LAMP / CRISPR-Cas test, considering RT-qPCR as the “gold standard”.

**Table 9:** Results of analytical and clinical (field) evaluations using the RT-qPCR and RT-LAMP / CRISPR-Cas tests. The degree of concordance is shown using the Kappa-Cohen index considering a 95% confidence interval (95% CI).

| RT-LAMP/CRISPR-Cas |  | RT-qPCR |  | Total | Kappa (CI 95%; p value) |
| --- | --- | --- | --- | --- | --- |
|  |  | Positive | Negative |  |  |
| Analytical Validation | Positive | 39 | 0 | 39 | 1.0 (1.0-1.0; p<0.00001) |
|  | Negative | 0 | 59 | 59 |  |
|  | Total | 39 | 59 | 98 |  |
| Clinical Validation (General) | Positive | 76 | 2 | 78 | 0.94 (0.89-0.98; p<0.00001) |
|  | Negative | 5 | 193 | 198 |  |
|  | Total | 81 | 195 | 276 |  |
| Clinical validation (Week 1) | Positive | 57 | 2 | 59 | 0.92 (0.87-0.98; p<0.00001) |
|  | Negative | 5 | 186 | 191 |  |
|  | Total | 62 | 188 | 250 |  |
| Clinical validation (Week 2) | Positive | 19 | 0 | 19 | 1.0 (1.0-1.0; p<0.00001) |
|  | Negative | 0 | 7 | 7 |  |
|  | Total | 19 | 7 | 26 |  |

**Figure 5.**
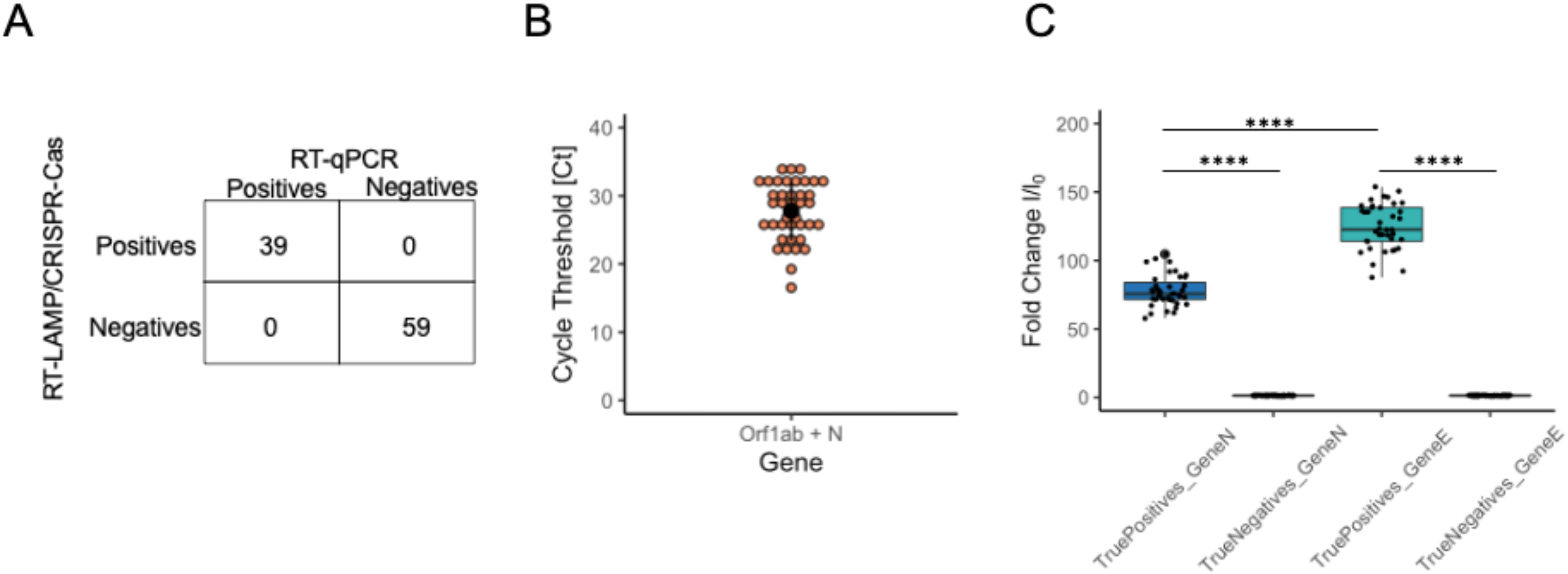
Retrospective analytical validation. The diagnostic performance of DETECTR was evaluated against RT-qPCR using 100 nasopharyngeal swabs from Peruvian patients, in triplicate and in blind. (A) 39 positive and 59 negative results were obtained with both methods, without false positives or false negatives. (B) Dot plot showing the cycle threshold values (Ct) of RT-qPCR for viral genes N and Orf1ab detected together in the same fluorescence channel. The horizontal line represents the arithmetic mean of all values with standard deviation. (C) Dot plot showing the fold change in fluorescence (y axis) for samples with the following DETECTR results: true positives for the E gene, true negatives for the E gene, true positives the N gene, and true negatives for the N gene (x axis). Fold change values (I/I_0_) were obtained by normalizing the fluorescent signal of the sample end-point (t=10 min) against the background signal at t=0. **** = p<0.0001 using the Kruskal-Wallis test, corrected with Dunn’s test, comparing all groups against the group of “true negatives” as a control group. Note that the fold change in fluorescence is significantly greater for the viral gene E compared to N.

### Clinical validation of RCSMS at two hospitals in Lima

Next, we set out to validate the field performance of our RCSMS procedure, which combines the biosensing capabilities of DETECTR (RT-LAMP/CRISPR-Cas12a coupled reactions) with the rapid and low-cost preparation of viral RNA with the TCEP/EDTA thermochemical method. The cohort reported here so far amounted to 276 individuals (current status from a total of 350 samples). Their distributions according to gender, age group, time of illness and symptoms are shown in Table 8. Interestingly, we noticed a higher proportion of female participants (n=158, 57.2%) and young adults (n=207, 75%), with a mean age of 44.5 years and standard deviation of 13.2. The dominant time of illness in the sample was 1 week of symptoms (n=242, 87.7%) and the mean time of symptoms was 4.1 days with a standard deviation of 2.5. Among the symptoms reported, sore throat predominated (n=181, 65.6%), followed by headaches (n=118, 42.8%). Ten individuals (3.6% of the total) did not record their date of symptom onset; therefore, their results were excluded from the analysis stratified by time of illness.

**Table 8:** Clinical and epidemiological data of the samples used for clinical validation. Of 276 individuals, their distributions according to gender, age group, time of illness and symptoms are shown.

| Sample characteristic |  | Quantity (n) | Percentage (%) |
| --- | --- | --- | --- |
| Sex | Male | 118 | 42.8 |
|  | Female | 158 | 57.2 |
| Age group | Young | 33 | 12.0 |
|  | Young Adult | 207 | 75.0 |
|  | Elderly | 34 | 12.3 |
| Sick time | 1 week | 242 | 87.7 |
|  | 2 weeks | 23 | 8.3 |
|  | Not registered | 10 | 3.6 |
| Symptoms | Throat pain | 181 | 65.6 |
|  | Fever | 91 | 33.0 |
|  | Headache | 118 | 42.8 |
|  | Anosmia | 19 | 6.9 |
|  | Nasal congestion | 53 | 19.2 |
|  | General discomfort | 97 | 35.1 |
|  | Diarrhea | 52 | 18.8 |
|  | Dysgeusia | 12 | 4.3 |
|  | Chest pain | 11 | 4.0 |
|  | Cough | 38 | 13.8 |
| RT-qPCR Result | Positive | 81 | 29.3 |
|  | Negative | 195 | 70.7 |
| RT-LAMP / CRISPR-Cas Result | Positive | 78 | 28.3 |
|  | Negative | 198 | 71.7 |

Of the 276 samples processed for detection, 78 gave positive results for the E gene by RCSMS (positivity = 28.3%) whereas 81 gave positive results by RT-qPCR (positivity = 29.3%) (Fig. 6). Statistical analyses (Table 7 and Table 9) yielded a prevalence of 29% (95% CI: 24.0-35.1), with a high concordance between the two tests (Cohen’s Kappa coefficient of 0.96; 95% CI: 0.94-0.99; p <0.00001), 93.8% sensitivity (CI 95%: 86.2-98.0), 99.0% specificity (CI 95%: 96.3-99.9), PPV of 97.4% (CI 95%: 91.0-99.7), NPV of 97.5% (95% CI: 94.2-99.2) and the Area Under the Curve (AUC) of the receiver operating characteristic (ROC) curve of 0.964 (95% CI: 0.937-0.991). In total, seven incongruencies (2.5%) were recorded between the two diagnostic tests: two false positives (false positive rate, FPR=2.6%) and five false negatives (false negative rate, FNR=2.5%). To assess the performance of RCSMS under prevalence conditions different from those found in this study, we carried out a statistical simulation of PPV and NPV un, assuming known and fixed prevalence values in all scenarios. The simulation predicts a very good tolerance of RCSMS to changes in prevalence, with NPV values >95% in prevalence settings of up to 50%, and PPV values >90% in prevalence settings >10% (Fig. 7).

**Figure 6.**
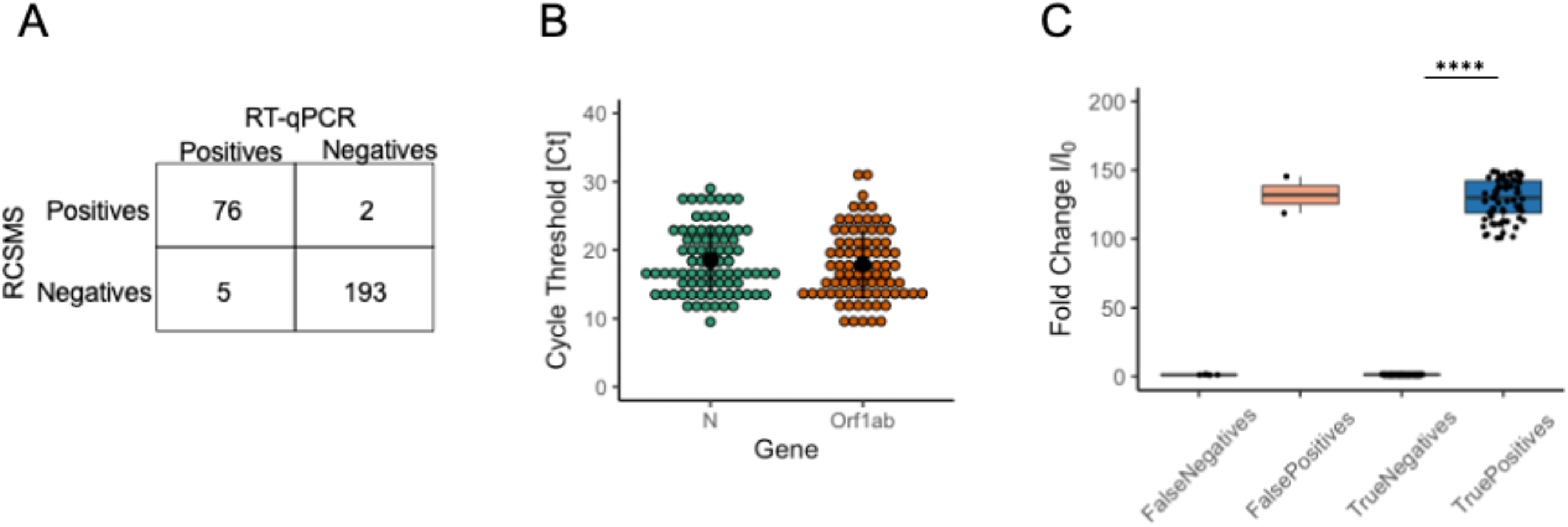
Clinical validation in two hospitals in Lima. The diagnostic performance of RCSMS was evaluated using 276 saliva samples from symptomatic patients at Guillermo Almenara and Edgardo Rebagliati Martins National Hospitals. Comparison was run against RT-qPCR data obtained with nasopharyngeal swab RNAs from the same patients. (A) RT-qPCR produced 81 positive and 195 negative results, whereas RCSMS yielded 78 positive results, with five false negatives and two false positives (relative to RT-qPCR). (B) Dot plot showing the cycle threshold values (Ct) of RT-qPCR for viral genes N and Orf1ab detected in separate fluorescence channels. The horizontal line represents the arithmetic mean of all values with standard deviation. (C) Dot plot showing the fold change in fluorescence (y axis) for samples with the following RCSMS results for the E gene: true positives, true negatives, false positives, and false negatives (x axis). Fold change values (I/I_0_) were obtained by normalizing the fluorescent signal of the sample end-point (t=10 min) against the background signal at t=0. **** = p<0.0001 using the Mann-Whitney U test for comparison of true positives with true negatives.

**Figure 7.**
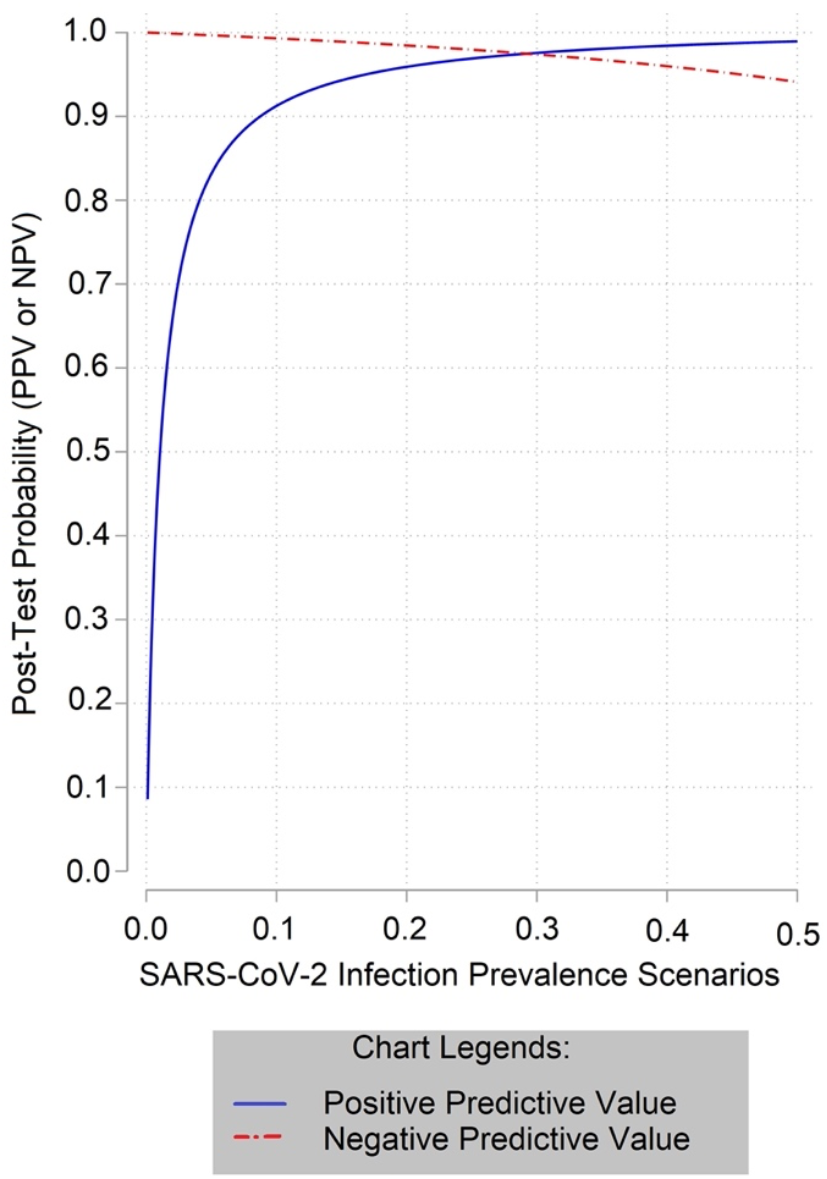
Post-test probability scenario simulations for PPV and NPV according to prevalences of SARS-CoV-2 infection. The graph shows the expected variation of the positive predictive values (PPV) and negative predictive values (NPV) for RCSMS according to different disease prevalence scenarios. X axis: prevalence of SARS-CoV-2 infection; y axis: PPV (blue line) and NPV (red line). In a scenario of >15% prevalence, both PPV and NPV >95%, whereas above 5% prevalence, the PPV >85%.

To assess whether the duration of symptoms affected the performance of the test, the same statistical analysis was performed, stratifying the data by week 1 and 2 of symptoms. On one hand, for the 242 patients in the first week of symptoms, a prevalence of 25% was obtained (95% CI: 20.0-30.6), with a Cohen’s Kappa coefficient between diagnostic tests of 0.924 (95% CI: 0.87-0.98; p <0.00001), 91.9% sensitivity (95% CI: 82.2-97.3), 98.9% specificity (95% CI: 96.2-99.9), PPV of 96.6% (95% CI: 88.3-99.6), NPV of 97.4% (95% CI: 94.0-99.1) and the Area Under the Curve (AUC) of the receiver operating characteristic (ROC) curve of 0.954 (95% CI: 0.919-0.989). In addition, seven incongruencies (2.8%) were recorded between the two tests: two false positives (TFP=2.6%) and five false negatives (TFN=2.5%). On the other hand, for the 26 patients in the second week of symptoms, a prevalence of 73% (95% CI: 52.0-88.0) was observed, with a Cohen’s Kappa coefficient between diagnostic tests of 1.0 (95% CI: 1.0-1.0; p <0.00001), 100% sensitivity (95% CI: 82.0-100.0), 100% specificity (95% CI:59.0-100.0), PPV of 100% (95% CI: 82.0-100.0), 100% NPV (95% CI: 59.0-100.0) and the Area Under the Curve (AUC) of the receiver operating characteristic (ROC) curve of 1.0 (95% CI: 1.0-1.0). In the second week, no false positives or false negatives were recorded, possibly due to the low representation of second-week patients in our study (most patients visit the hospital during the first week of symptoms). This stratified analysis suggests a potential increase of 3.4% in sensitivity and 2.6% in specificity of our RCSMS test between the first and second week of disease. Finally, by using serial dilutions of synthetic RNA from 5,000 to 1 viral copy per 10 µl RT-LAMP reaction, we found the limit of detection for our method (either using fluorescence or immunochromatography) to be 5 viral copies per RT-LAMP reaction of 10 µl (Fig. 4C and Table 5), which is in close agreement with the limits of detection reported for DETECTR (8) and RT-qPCR (11-13).

## DISCUSSION

Perú ranks consistently at the top among the countries most affected by COVID-19 worldwide. Throughout 2020, the country faced limited testing capacity, inadequate public health services and the lack of local biotechnology. As of April 2021, Perú is in the midst of a second pandemic wave, worsened by the surge of more infectious and deadly viral variants, highlighting the urgent need to make COVID-19 testing available to urban and rural settings with limited access to state-of-the-art diagnostic laboratories.

The present study describes the implementation and clinical validation in Peruvian patients of RCSMS, a DETECTR-based molecular test capable of detecting SARS-CoV-2 genes from saliva in ∼40 minutes, with high sensitivity and specificity, without sophisticated instrumentation, specialized personnel or expensive laboratory supplies, such as transport media and RNA extraction kits. When comparing the diagnostic performance of RCSM vs RT-qPCR, we found an excellent level of agreement between the efficiency of the two tests, both under controlled conditions in the laboratory (Cohen’s Kappa coefficient = 1.00) and with clinical samples in the field (Cohen’s Kappa coefficient = 0.96). These values are consistent with the performance of other molecular tests that successfully use CRISPR-Cas technology for the detection of SARS-CoV-2 from nasopharyngeal swabs (8, 17, 18). Given that the analytical and clinical validations used the exact same experimental protocols and laboratory settings, we interpret this slight reduction in Cohen’s Kappa coefficient, from 1.00 to 0.96 as a result of the intrinsic variability expected from sample collection and handling procedures under field conditions.

To this date, we are not aware of field validation reports for other saliva, CRISPR-Cas-based tests for SARS-CoV-2 detection. Although nasopharyngeal swabs are a standard procedure for the collection of respiratory virus samples (3), their use raises the cost of testing, causes discomfort to the patient and increases the probability of transmission to and among sample collection personnel. Therefore, saliva tests are a significant step towards making COVID-19 diagnostics a routine task. Importantly, RCSMS implementation does not require biosafety level 2 laboratories because the lysis and inactivation of viral particles occur immediately after saliva collection, turning samples non-infectious and keeping them safe from degradation by nucleases. Furthermore, sample stability is an important precondition for the development of high-quality, point-of-care tests.

LAMP is a simple, robust and low-cost DNA amplification method that can be visualized by absorbance, gel electrophoresis or colorimetric reactions (19, 20). However, these readouts do not discriminate between specific products and those obtained from the artifactual amplification of complex or contaminated templates (21). Because LAMP is a highly efficient reaction, diagnostic methods consisting only of this step need careful handling to prevent aerosol contamination. In our hands, this was achieved by setting up LAMP reactions in a designated work area, separate from all other steps of the RCSMS protocol, and keeping the reaction tubes closed until used in the detection reaction. Notably, the CRISPR-Cas12a system adds precision to the protocol because it produces a signal only when the viral sequence is specifically recognized and cut by the Cas12a enzyme. This reaction sets off the indiscriminate cut -also by Cas12a- of hundreds of reporter molecules, thus amplifying the original LAMP signal with high resolution and intensity (22).

In order to physically recognize nucleic acids, Cas12a must be “guided” by an RNA molecule complementary to its target sequence. This guide RNA has a bipartite structure, with a “stem” that forms a complex with Cas12a and a leader that matches and binds the target DNA sequence, giving the system its specificity (23-27). The optimal size and sequence of guide RNAs vary according to the type of Cas enzyme and the target sequence to be detected. For instance, for Cas9 -the first endonuclease developed for genome editing (23)-the guide stem is 80 bp long and the leader varies between 17-23 bp (24), whereas Cas12a requires a 20 bp stem and a 21-23 bp leader (25, 26), and Cas13a -which recognizes RNA targets-uses a 36 bp stem and a 28 bp leader (27). Therefore, the design of guide RNAs using bioinformatic tools is crucial for the diagnostic specificity of RCSMS and all other CRISPR-Cas-based biosensors. The guide RNAs used here (41 bp) efficiently detected the viral sequences targeted by Broughton and colleagues in their DETECTR study with E and N genes (8). Similar to what they report in their analytical validation, we observed more efficient and consistent detection of the E gene. This was particularly evident from the fluorescence signal levels (Fig. 5C) and kinetics (Fig. 3). Conversely, Brandsma et al. clinically validated the same system in three Dutch hospitals, but targeting only the N gene (17). Interestingly, a clinical validation study by Patchsung et al. using a Cas13-based system found that the S gene (encoding the Spike protein) performed better than the Orf1ab and N genes (18). We also attempted detection of the S gene but obtained only spurious and highly variable results (data not shown). This may be explained by the fact that their system used a different enzyme (Cas13a) and guide RNAs to detect different target sequences on a different type of nucleic acid (*in vitro* transcribed RNA, post-amplification). Given that the Spike protein mediates cellular entry of the virus, it is also relevant to consider the role of positive selection on the fixation of S gene mutations associated with infectivity and immunoevasion, as seen in the P.1 (“Brazilian”), B.1.351 (“South African”), B.1.1.7 (“British”) variants (28-30). For example, samples from a large number of UK COVID-19 patients were shown to have S genes that could not be amplified by RT-qPCR due to the Δ69/Δ70 and N501Y mutations of B.1.1.7 (29, 30). Interestingly, patients with this “*S gene target failure*” (SGTF) had a 55% higher risk of death than the original Wuhan strain (29, 30), confirming that the B.1.1.7 is more transmissible and lethal than pre-existing SARS-CoV-2 variants. For diagnostic purposes, it is therefore advisable to target more conserved genetic sequences. Nonetheless, the versatility and specificity of DETECTR allow for rapid reprogramming of RCSMS (28), simply by redesigning primers and guide RNAs to target new variant sequences.

The rapid evolution of SARS-CoV-2 genomes makes it necessary to continuously screen the population for virus variants to ensure that the performance of molecular tests is not affected by mutations. Our bioinformatic analysis of 944 SARS-CoV-2 genomes from Peruvian patients revealed the presence of a small number of mutations in one primer for the E gene and one primer for the N gene, out of a total of twelve primers evaluated. However, these point mutations occur at low frequency in the population and did not significantly affect DNA amplification, as evidenced by the high sensitivity and specificity values obtained here for RCSMS. While we had no access to clinical samples containing other human coronaviruses, the ability of DETECTR to detect the E and N genes of SARS-CoV-2 (and not those of other related human coronaviruses) has been demonstrated *in silico* and *in vitro* by Broughton et al (8). Furthermore, we found that the performance of RCSMS was not affected by minor variations in experimental conditions. For instance, 10 µl LAMP reactions performed efficiently between 60 and 65°C and CRISPR-Cas detection was not compromised when using half the concentration of Cas12a or guide RNAs, reducing the reaction time to 5 min, or varying the input DNA volume between 0.2 and 3 µl. Similarly, a 6-fold increase in the concentration of fluorescent probes or a 0.1X variation in the concentration of biotinylated probes had no appreciable effect on detection.

Using 100 nasopharyngeal swabs from Peruvian patients, DETECTR performed indistinguishably from RT-qPCR under controlled laboratory conditions, reproducibly reaching relative values of 100% of specificity and sensitivity, and a LOD of one viral copy per 10 µl RT-LAMP reaction. These data are in agreement with the excellent results obtained in similar validations done in the Netherlands and the USA (8, 17). The field validation with 276 patients at Almenara and Rebagliati National Hospitals further highlights the remarkable performance of our saliva-based RCSMS test against conventional RT-qPCR from nasopharyngeal swabs, yielding 93.8% sensitivity and 99.0% specificity values. We are not aware of other field validated saliva tests that use CRISPR-Cas technology; nonetheless these results are very similar to those reported for DETECTR by Broughton et al (sensitivity 95% and specificity 100% in analytical validation) (8) and Brandsma et al (sensitivity 95.5% and specificity 92.9% in clinical validation) (17), and for the Cas13 test by Zhang’s group (sensitivity 96% and specificity 100% in clinical validation) (18). Moreover, while we found seven incongruencies (two false positives and five false negatives) in 276 samples, Brandsma et al. found 21 (10 false positives and 11 false negatives) in 378 samples (16), and Patchsung et al. reported ten false negatives in 154 samples (18). In these studies, the LOD from nasopharyngeal swabs reached 50 and 42 copies per 25 µl reaction (17, 18). Notably, RCSMS routinely detected 50 copies per 10 µl reaction in triplicate assays, occasionally reaching five copies in one or two replicates.

Sensitivity and specificity are inherent properties of a test and therefore do not change between populations. However, the usefulness of a test, inferred from its predictive value, varies according to the true prevalence of the disease in a population. This is particularly relevant for COVID-19, where prevalence follows the evolution of complex geographical and temporal patterns of transmission, owing to demographic structure, human travel and migration. We estimated the predictive value of a test under different prevalence settings to determine adequate scenarios for its use. Generally, the NPV of diagnostic tests is expected to drop in high prevalence settings, as the number of false negatives increase (31). Nonetheless, our statistical simulation for RCSMS using the currently available data (276 out of 350 samples analyzed, Fig. 7), predicts excellent NPVs >95% for prevalence scenarios as low as 1% and as high as 50%. Ensuring a low rate of false negatives (positive individuals being deemed negative) is especially desirable considering that COVID-19 is a serious, largely asymptomatic and contagious disease, for which early measures are advisable. Thus, a negative RCSMS result would maintain a very good level of accuracy in high prevalence settings (i.e. at a hospital with a high proportion of COVID-19 patients), even as the prevalence sinks due to the natural progression of the epidemic curve. Moreover, these data indicate that RCSMS could be used with confidence to rule out infection in low prevalence settings such as airports and land travel facilities, schools, universities, sporting or cultural events, as well as religious services and working environments with high flow of personnel, where ensuring negativity is important.

Our simulation also obtains adequate PPVs >95% and >90% for prevalence scenarios >20% and 10%, respectively. These PPV estimates are consistent with the results from our field study, where prevalence reached almost 30%, strengthening the conclusion that RCSMS is particularly well suited for the identification of positive cases during acute pandemic phases, such as the current second waves hitting Perú, India and Brazil. A sharp decrease of PPV in low prevalence settings is common to all diagnostic tests because of the higher probability of obtaining false positives (31). Notably, according to our simulation, RCSMS is expected to perform reasonably well (PPV>85%) even at a 5% prevalence scenario. As the pandemic recedes and the COVID-19 prevalence in the population decreases below 5%, it will be advisable to complement positive results using other molecular tests as well as diagnostic criteria (symptoms, oxygen saturation, chest X-ray/CT scan). Altogether, our data strongly suggest that RCSMS is sensitive enough to avoid missing infected individuals in high prevalence settings, and specific enough to maintain a very low proportion of erroneously diagnosed cases in low prevalence settings.

In conclusion, the present study integrates two methodologies that had been separately applied to COVID-19 testing: 1) the use of inactivated saliva as starting material for viral gene amplification (32) and 2) the use of DETECTR as a SARS-CoV-2 biosensor (8). The resulting molecular test, RCSMS, can be easily implemented at the primary care level and diagnostic laboratories alike, using a simple lateral flow readout. To our knowledge, this is the first *clinical validation* of a CRISPR-Cas-based test for COVID-19 in Latin America using saliva, and one of the few of its type developed worldwide (33, 34, 35). Noteworthy, an Argentinian group recently reported a DETECTR system (36) that uses nasopharyngeal swabs and fluorescence readouts. Within this group of newly validated CRISPR-Cas-based tests for the detection of SARS-CoV-2, RCSMS has the potential of becoming an important tool for the massification of COVID-19 molecular tests in Peru and neighboring countries.

## Supporting information

RCSMS detection with lateral flow strips

## Data Availability

Data available upon request

https://drive.google.com/file/d/1yNYNYKWukbyAGIF2kK-jogDfUGn4xPT2/view

## CONTRIBUTIONS

J.A.N., A.P.O., P.R.A., J.B.L., R.G.L. and E.M.T designed and planned the project. J.A.N., J.B.L., R.G.L. and E.M.T implemented the molecular method. J.A.N., J.B.L., R.G.L., A.Q.R. and E.M.T carried out the analytical validation. J.L.M., J.A.N and E.M.T. designed the clinical validation. J.L.M., E.R.C., F.A.L.M., J.A.N., A.Q.R. and E.M.T. planned, coordinated and executed field work in hospitals as well as RT-qPCR analysis for clinical validation. J.A.N., A.Q.R., J.L., P.R.A., and E.M.T. performed RCSMS analysis for clinical validation. All authors contributed to the analysis of different aspects of the data. J.A.N., P.R.A., J.L., J.L.M. and E.M.T wrote the article.

## ACKNOWLEDGEMENTS

We are profoundly indebted to: C.M. Caro, A. Rozas, J. Arriola, H. Añaños, J.C., F. León-Velarde, A. Sobarzo and especially M. and E. Arias for their generous support to this project, from the search for funding sources to assistance with the resolution of regulatory and bureaucratic hurdles. C. Guerra for providing personnel, equipment and reagents, as well as support with administrative procedures. P. Soto for critical reading of the manuscript. T. Ochoa and D. Durand for the use of their laboratory equipment. P. Tsukayama for his help with bioinformatic analyses. V. Adaui and P. Milón for helpful technical discussions. F. Molinelli, E. Carlín, L. Shica, J. Santillana, J. Amorós, P. Pimentel, C. Bedoya, G. Minaya, Y. Hurtado, R. Araujo, G. Solis and C. Díaz-Vélez from ESSALUD, for organizing and facilitating the collaboration with their hospital network for field studies. K. Valverde, C. Morales, H. Pinedo and R. Álvarez for their valuable work in collecting saliva samples from the participants. The local and international press that helped bring attention and support to this project. Merck del Perú, GenLab del Perú, AC Interlab, La Ensenada, Mased and Ahseco for granting priority access to supplies and equipment. We especially want to express our deepest gratitude to the symptomatic patients who attended the COVID-19 triage areas of Guillermo Almenara and Edgardo Rebagliati National Hospitals, for their generous and selfless participation in this study, despite their precarious condition and the often pressing circumstances; for their words of encouragement towards Peruvian science, and for their great humanity. We dedicate our work to them and their families, hoping that they were able to overcome the disease. Finally, our deep condolences to all those who have lost loved ones during this pandemic.

